# Development of a Deep Learning Model Integrating CT Images and Blood Data for the Diagnosis of Acute Cholecystitis

**DOI:** 10.64898/2026.05.08.26352724

**Authors:** Takeshi Horaguchi, Ryosuke Nomura, Shunsuke A Sakai, Nao Saito, Kyoko Kurihara, Masaki Ohira, Ritsuki Takaha, Noriki Mitsui, Yuji Hatanaka, Ryoma Yokoi, Masashi Kuno, Hirokatsu Hayashi, Masahiro Fukada, Yuta Sato, Itaru Yasufuku, Ryuichi Asai, Hideaki Bando, Riu Yamashita, Nobuhisa Matsuhashi

## Abstract

**Purpose:** In this study, we aimed to develop and evaluate an artificial intelligence-based diagnostic model for the diagnosis of acute cholecystitis (AC) using non-contrast CT images and clinical data.

**Materials and Methods:** This retrospective study included 199 patients (100 AC, 99 non-AC) treated between January 2016 and December 2025 at a single center. Patients were randomly divided into training (n=139) and test (n=60) datasets. Three models were constructed: an imaging-based deep learning model, a clinical data-based machine learning model, and a hybrid machine learning model integrating deep learning-derived imaging features with clinical data. CT images were preprocessed, and gallbladder regions were segmented. Clinical variables included white blood cell counts and levels of C-reactive protein and liver function markers. Model performance was evaluated using accuracy, precision, recall, specificity, F1 score, and area under the receiver operating characteristic curve (AUC). Statistical comparisons were performed using Welch’s t-test and Chi-square test.

**Results:** The imaging-based model achieved accuracy 0.883, precision 0.848, recall 0.933, specificity 0.833, and AUC 0.916. The blood-based model achieved accuracy 0.917, precision 0.931, recall 0.900, specificity 0.933, and AUC 0.949. The hybrid model showed the highest performance, with accuracy 0.950, precision 0.909, recall 1.000, specificity 0.900, F1 score 0.952, and AUC 0.986.

**Conclusion:** A hybrid model integrating CT imaging and clinical data improved diagnostic performance for AC compared with single-modality models.

## Introduction

Acute cholecystitis (AC) is a leading cause of acute abdomen in emergency departments, accounting for 3–10% of patients presenting with abdominal pain (1,2). Rapid progression can lead to severe complications, such as gangrenous cholecystitis, perforation, abscess formation, and sepsis, necessitating early diagnosis and immediate therapeutic decision-making (3). The Tokyo Guidelines 2018 provide a widely accepted framework for diagnosis and severity grading by integrating local signs, systemic inflammation, and imaging features (1). While the Tokyo Guidelines 2018 criteria demonstrate high sensitivity and specificity, their implementation remains contingent on the quality of clinical and radiological assessments.

Ultrasonography is an established first-line imaging modality for acute abdomen due to its non-invasive nature, accessibility, and real-time utility (4). However, its diagnostic performance is highly operator-dependent and constrained by factors such as examiner experience, patient body habitus, and bowel gas (5). In emergency settings, where initial assessments are often performed by junior residents or non-specialists, this variability poses a significant challenge to diagnostic consistency. In contrast, computed tomography (CT) offers more objective and comprehensive evaluation, facilitating the detection of gallbladder inflammation, complications, and alternative causes of acute abdomen (6). Recent studies have indicated that CT has diagnostic performance comparable to that of ultrasonography, with a sensitivity and specificity of 83.9% and 94.0%, respectively, compared to 79.0% and 93.6% for ultrasonography (7).

In clinical practice, contrast-enhanced CT is frequently contraindicated by contrast medium allergy or renal dysfunction, rendering non-contrast CT the default initial imaging modality. This is particularly relevant in emergency settings, where time constraints necessitate rapid acquisition. However, non-contrast CT exhibits limited sensitivity for detecting AC, especially in early or suppurative cases, highlighting the need for enhanced diagnostic approaches (6,8).

Recent advances in artificial intelligence, particularly with deep learning-based models, have shown significant potential in medical image analysis by extracting subtle features that are not easily detectable by humans (9–13). In acute abdominal imaging, artificial intelligence, including deep learning approaches, can improve diagnostic accuracy, reduce interpretation time, and support triage (14). While deep learning-based models using ultrasonographic images have been reported, these rely on operator-acquired or pre-selected images, which introduces selection bias and limits applicability in real-world settings (15). Conversely, CT protocols are relatively standardized, making them ideal for automated artificial intelligence-based analysis (16,17). In particular, recent approaches that decompose Hounsfield units (HU) into three RGB channels for model input have been shown to improve the predictive performance of CT-based analyses (18). Furthermore, the clinical diagnosis of AC routinely incorporates laboratory data, such as elevated levels of inflammatory markers, suggesting that a multimodal approach integrating imaging with clinical metadata may further enhance diagnostic performance.

In this study, we developed and evaluated a deep learning-based imaging model and complementary machine learning models for diagnosing AC using non-contrast CT images, emphasizing real-world applicability. Additionally, we constructed a hybrid machine learning model incorporating blood test data with deep learning-derived imaging features and assessed its interpretability using visualization techniques. Finally, we explored a sensitivity-oriented deployment strategy to support triage by junior physicians and non-specialists in emergency care settings.

## Materials and Methods

### Study Population and Ethics

The study protocol was approved by the Institutional Review Board of Gifu University Hospital (Approval No. 2024-007). This retrospective cohort study was conducted at Gifu University Hospital, spanning cases from January 2016 to December 2025. Patients were identified through a retrospective review of consecutive clinical records. The exclusion criteria included incomplete clinical/laboratory data or suboptimal CT image quality (e.g., severe motion artifacts). A total of 199 patients were enrolled, comprising 100 patients with AC and 99 control participants without AC. The diagnosis of AC was established in accordance with the Tokyo Guidelines 2018 criteria, integrating clinical symptoms, laboratory markers, and imaging findings. Surgical pathology and/or clinical follow-up served as the reference standards. All CT images had corresponding radiology reports. For each case, two gastrointestinal surgeons (T.R., with 7 years of clinical experience, and T.H., with 12 years of clinical experience), both board-certified surgeons, independently selected a single representative axial CT slice. The representative image was defined as the slice considered most morphologically informative for gallbladder evaluation. In most cases, this corresponded to the slice showing the largest gallbladder diameter. In non-AC cases, the slice showing the largest visible gallbladder was similarly selected. When the two reviewers disagreed, the final slice was determined by consensus. Biochemical data were extracted from electronic medical records at the time of CT imaging.

### Data Preprocessing and Image Transformation

DICOM images were processed using the pydicom library (version 3.0.1). Raw pixel values were converted to HU using rescale slope and intercept parameters. To enhance soft-tissue contrast, images were windowed to a range of −70 to 170 HU, followed by normalization to 8-bit grayscale.

To leverage the transfer learning capabilities of models pretrained on natural images, we generated three-channel pseudo-color RGB images by applying three distinct HU window settings (18): Red (R), −70 to 50 HU; Green (G), −10 to 110 HU; and Blue (B), 50 to 170 HU. The resulting images were saved in PNG format for model input.

White saturation (blowout) artifacts were corrected using a rule-based image processing approach implemented with OpenCV (version 4.13.0.92) and NumPy (version 1.26.4). First, candidate pixels for white saturation were identified based on intensity and color information. Specifically, pixels with a mean RGB intensity ≥ 210 were selected, as well as pixels with a value (V) ≥ 210 and saturation (S) ≤ 70 in the HSV color space. Next, connected component analysis with 8-connectivity was applied to the binary mask, and only components with an area ≥ 10 pixels were retained. To refine the detected regions, morphological closing was performed using an elliptical kernel (7 × 7), followed by dilation using an elliptical kernel (5 × 5) for two iterations. For each detected region, a surrounding ring-shaped area was defined by subtracting the original mask from its dilated version. The pixel values within the white saturation regions were then replaced by the median RGB values computed from the corresponding ring regions, enabling local intensity interpolation while preserving surrounding structural information.

### Gallbladder Segmentation and Region of Interest Localization

Automated gallbladder segmentation was performed using TotalSegmentator (19) (version 2.12.0). A bounding box was generated from the segmentation mask, incorporating a 15-pixel margin in all directions to capture the pericholecystic environment. In four cases where automated segmentation was suboptimal, manual correction was performed by a specialist to ensure the gallbladder remained the central region of interest.

To mitigate the impact of imaging artifacts, overexposed regions were corrected using an intensity-based thresholding algorithm. Pixels were identified as artifacts if the mean RGB intensity was ≥ 210 or if the Value (V) was ≥ 210 with a Saturation (S) ≤ 70 in the HSV color space. These regions were refined via morphological closing and dilation, and subsequently inpainted using the median RGB values of the surrounding tissue.

### Deep Learning Framework

Input images were resized to 128 × 128 pixels and converted into three-channel (RGB) format. The imaging model employed an EfficientNet-B0 backbone pretrained on ImageNet, implemented using TensorFlow (version 2.16.2). The architecture included a Global Average Pooling layer, a fully connected (dense) layer with 128 units (GELU activation), a dropout layer (rate = 0.4), and a final sigmoid output layer for binary classification.

Training was executed in two phases:

1. Warm-up: The backbone was frozen, and the classification head was trained using the AdamW optimizer (learning rate = 1 × 10^-4^)
2. Fine-tuning: The final 10 layers of the backbone were unfrozen and optimized using Adam (learning rate =1 × 10^-5^), while Batch Normalization layers remained frozen to preserve internal covariate shift stability

Training was conducted with a batch size of 32 for up to 300 epochs, utilizing early stopping (patience = 10) based on validation loss. Following the training phase, this model served as a feature extractor: the 128-dimensional latent vectors from the penultimate dense layer were extracted and utilized as input features for the subsequent downstream machine learning models.

### Model Architecture and Training

We developed and compared three diagnostic frameworks: the Imaging Model (a deep learning model using CT data); the Blood Model (a machine learning model using blood data); and the Hybrid Model (a machine learning model integrating deep learning-derived imaging features and clinical data). The dataset was partitioned at the patient level into a training set (70%, n = 139) and a test set (30%, n = 60) to prevent data leakage. Automated machine learning was implemented via the PyCaret library (version 3.3.2) to identify the optimal classification architecture. The input feature space consisted of standardized biochemical laboratory data, the 128-dimensional latent vectors extracted from the deep learning backbone, or a concatenated vector of both for the hybrid approach.

To ensure a robust assessment of generalizability, model performance was rigorously evaluated on an independent test set. The primary metrics for discriminative capacity included accuracy, precision, recall (sensitivity), specificity, and F1 score. Additionally, the area under the receiver operating characteristic curve (AUC) was calculated to evaluate the overall diagnostic performance.

### Blood-Based and Hybrid Machine Learning Models

Clinical variables (white blood cell count, platelet count, levels of C-reactive protein, total bilirubin, aspartate aminotransferase, alanine aminotransferase, gamma-glutamyl transpeptidase, alkaline phosphatase, and creatinine, and the prothrombin time–international normalized ratio) were standardized using z-score normalization. For the Blood Model, an Extra Trees classifier was selected as the final estimator. For the Hybrid Model, 128-dimensional feature vectors extracted from the bottleneck layer of the deep learning model were reduced using principal component analysis, retaining components that explained more than 90% of the cumulative variance (n = 41; Figure S1). These features were then concatenated with the clinical variables, and a Random Forest classifier was used as the final estimator for the Hybrid Model. Model selection for both models was performed using PyCaret based on cross-validated performance.

### Statistical Analysis

Baseline characteristics were compared between the AC and non-AC groups using Welch’s two-sample t-test for continuous variables and the chi-square test for categorical variables. All statistical analyses were implemented in R statistical software (version 4.5.3), with a *p*-value < 0.05 considered statistically significant.

### Data Availability

All scripts used in this study are available on GitHub (https://github.com/RyosukeNomural/model_for_acute_cholecystitis).

## Results

### Patient Characteristics and Dataset Partitioning

A total of 199 patients (100 with AC and 99 without AC) were enrolled in this study (Table 1). Baseline demographic characteristics were generally comparable between the two groups, with no significant differences in sex, age, height, or serum creatinine levels. However, patients with AC had significantly higher body weight and body mass index than those without AC. Laboratory findings also differed significantly between the groups: the AC group had higher white blood cell counts, C-reactive protein levels, total bilirubin levels, liver enzyme levels (aspartate aminotransferase, alanine aminotransferase, gamma-glutamyl transpeptidase, and alkaline phosphatase), and the prothrombin time–international normalized ratio, while platelet counts were significantly lower.

**Table 1.** Demographic and clinical characteristics of the study cohort.

|  | Non-AC (n=99) | AC (n=100) | *p*-value |
| --- | --- | --- | --- |
| Sex |  |  | 0.121 |
| Male | 77 (77.8%) | 68 (68.0%) |  |
| Female | 22 (22.2%) | 32 (32.0%) |  |
| Age | 67.4 ± 12.6 | 66.3 ± 14.3 | 0.583 |
| Height [cm] | 163.7 ± 9.1 | 161.5 ± 9.6 | 0.100 |
| Weight [kg] | 55.1 ± 9.1 | 60.1 ± 11.1 | < 0.001 |
| BMI [kg / m<sup>2</sup>] | 20.6 ± 3.3 | 23.0 ± 3.6 | < 0.001 |
| WBC [ /μL] | 6709 ± 2331 | 12571 ± 5412 | < 0.001 |
| PLT [× 10<sup>4</sup>/μL] | 24.6 ± 8.3 | 21.6 ± 9.4 | 0.017 |
| CRP [mg/dL] | 0.662 ± 1.48 | 12.6 ± 10.5 | < 0.001 |
| T-Bil [mg/dL] | 0.80 ± 0.30 | 1.71 ± 1.2 | < 0.001 |
| AST [U/L] | 20.8 ± 10.0 | 75.9 ± 138.6 | < 0.001 |
| ALT [U/L] | 18.4 ± 14.5 | 63.3 ± 118.5 | < 0.001 |
| γGTP [U/L] | 44.3 ± 37.8 | 120.5 ± 236.4 | < 0.001 |
| ALP [U/L] | 95.2 ± 54.2 | 245.5 ± 236.4 | < 0.001 |
| Cre [mg/dL] | 0.933 ± 0.446 | 0.9494 ± 0.646 | 0.835 |
| PT-INR [-] | 0.951 ± 0.124 | 1.08 ± 0.232 | < 0.001 |

Data are presented as numbers for categorical variables and means ± standard deviations for continuous variables. AC, acute cholecystitis; ALP, alkaline phosphatase; ALT, alanine aminotransferase; AST, aspartate aminotransferase; BMI, body mass index; Cre, creatinine; CRP, C-reactive protein; PLT, platelet; PT-INR, prothrombin time-international normalized ratio; T-Bil, total bilirubin; WBC, white blood cell; γGTP, gamma-glutamyl transferase

### Image Processing and Feature Extraction

Three diagnostic architectures were developed and evaluated: the Imaging Model, the Blood Model, and the Hybrid Model integrating both data streams (Figure 1). To optimize deep learning performance, CT DICOM images underwent a standardized preprocessing pipeline. Raw data were converted to HU and normalized to 8-bit grayscale. To enhance feature representation, three-channel RGB inputs were generated using varying HU window settings. The gallbladder was segmented using TotalSegmentator, with regions of interest extracted including a pericholecystic margin; manual refinement was performed where necessary. These processed images served as input for EfficientNet, from which 128-dimensional latent feature vectors were extracted for downstream machine learning classification (Figure S2A–D).

**Figure 1.**
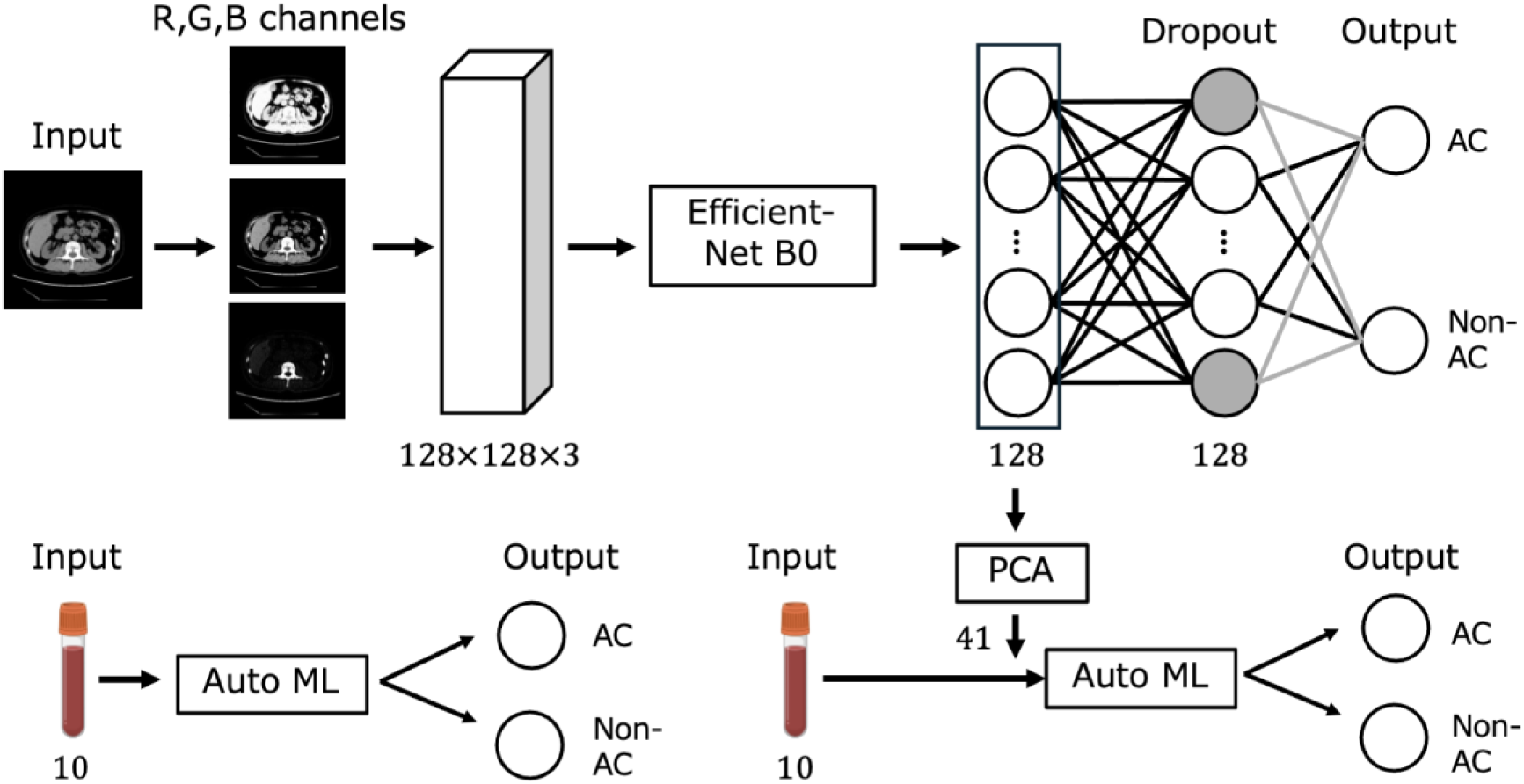
(A) Overview of Imaging model. Input CT images were converted into three-channel (RGB) format and resized to 128 × 128 × 3 pixels before being processed by an EfficientNet-B0–based deep learning model. The model generated a binary output for acute cholecystitis (AC) versus non-acute cholecystitis (non-AC). (B) Overview of Blood model. Ten clinical blood variables were used as input for a machine learning classifier to predict AC versus non-AC. (C) Overview of Hybrid model. A 128-dimensional latent feature vector extracted from the imaging model was reduced to 41 dimensions by principal component analysis (PCA) and combined with the 10 blood variables. The integrated features were then used as input for a machine learning classifier to predict AC versus non-AC.

### Performance of the Imaging Model

The Imaging Model, utilizing the 128-dimensional EfficientNet features, demonstrated stable convergence during training (Figure 2A, B). On the independent test dataset, the model yielded an accuracy of 0.883, precision of 0.848, recall of 0.933, specificity of 0.833, and an F1 score of 0.889 (Figure 2C). The AUC was 0.916 (Figure 2D). These metrics indicate robust discriminative performance, with high sensitivity (recall) highlighting the model’s potential to minimize missed diagnoses.

**Figure 2:**
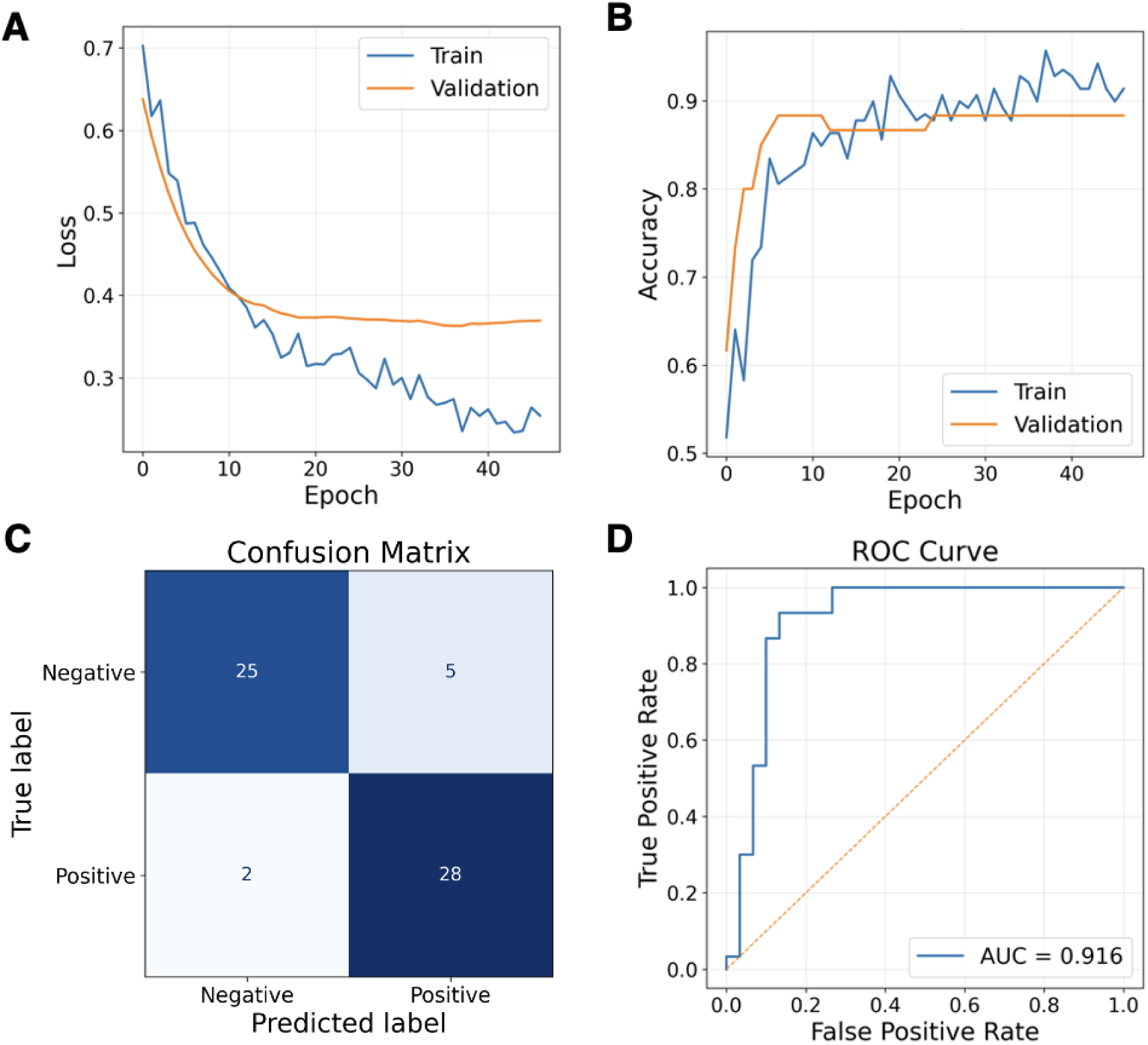
(A) Line graph showing the training and validation loss over epochs. (B) Line graph showing the training and validation accuracy over epochs. (C) Confusion matrix showing the true and predicted labels. (D) Receiver operating characteristic (ROC) curve evaluating the classification performance. The dashed line represents the performance of a random classifier. AUC = area under the curve.

Gradient-weighted Class Activation Mapping visualization confirmed that, in true-positive cases, the model consistently localized its attention to the gallbladder and its immediate anatomical environment (Figure S3A). In true-negative cases, minimal activation was observed in the gallbladder region, reflecting a lack of pathological morphological markers (Figure S3B). However, in misclassified cases (false positives and false negatives), attention maps were less localized, suggesting that relying solely on morphological features like gallbladder volume or wall thickness may be insufficient for definitive diagnosis in atypical presentations (Figure S3C, D).

### Performance of the Blood Model

The Blood Model integrated ten laboratory variables: white blood cell count, platelet count, levels of C-reactive protein, total bilirubin, aspartate aminotransferase, alanine aminotransferase, gamma-glutamyl transpeptidase, alkaline phosphatase, and creatinine, and the prothrombin time–international normalized ratio. Following an automated machine learning comparison using PyCaret, the Extra Trees Classifier was identified as the optimal architecture.

On the test dataset, this model achieved an accuracy of 0.917, precision of 0.931, recall of 0.900, specificity of 0.933, and an AUC of 0.949 (Figure 3A, B). Feature importance analysis identified the C-reactive protein level as the primary contributor, followed by the white blood cell count and total bilirubin level (Figure 3C), underscoring the centrality of systemic inflammatory biomarkers in the classification task.

**Figure 3:**
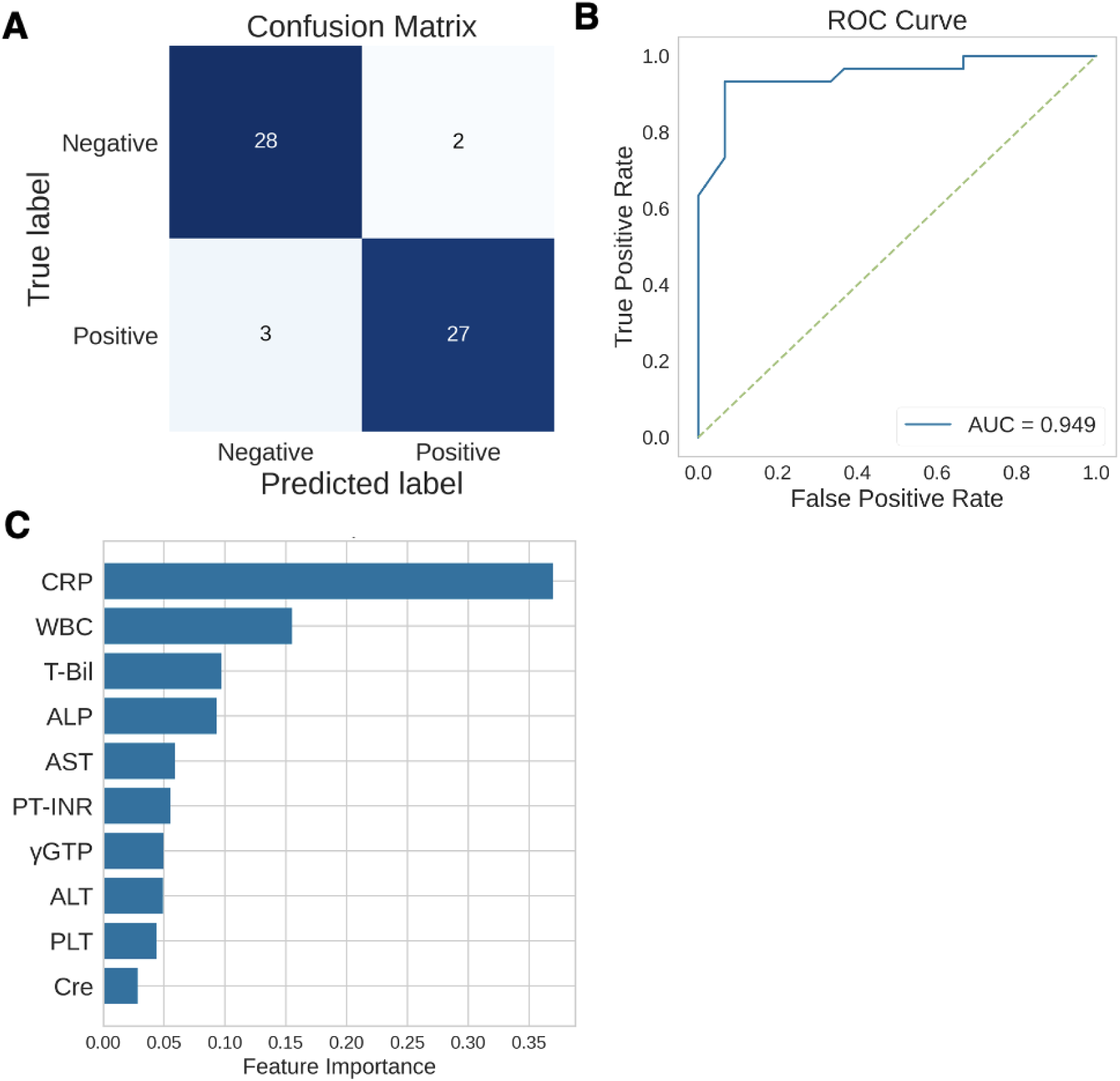
(A) Confusion matrix showing the true and predicted labels for the model. (B) Receiver operating characteristic (ROC) curve evaluating the classification performance. The dashed line represents the performance of a random classifier. (C) Bar chart displaying the relative feature importance of all laboratory variables contributing to the model’s predictions.

Analysis of five misclassified cases (three false negatives and two false positives) revealed that these instances lacked a clear correlation between clinical presentation and gallbladder morphology (Figure S3A, B). Notably, false-positive cases exhibited markedly elevated C-reactive protein levels (> 9 mg/dL) without anatomical AC evidence, while false-negative cases showed low C-reactive protein levels (< 1 mg/dL) despite potential clinical indicators (Table S1). This suggests the Blood Model is highly sensitive to inflammatory signals but lacks specificity in the absence of structural confirmation.

### Performance of the Hybrid Model

Finally, we developed a hybrid machine learning model using a Naive Bayes classifier to fuse imaging-derived features with clinical laboratory data. This integrated approach achieved superior performance on the test dataset, with an accuracy of 0.950, precision of 0.909, recall of 1.000, specificity of 0.900, and an F1 score of 0.952 (Figure 4A). The AUC reached 0.986 (Figure 4B). Feature importance analysis revealed that both imaging-derived and laboratory features contributed meaningfully to the model’s predictions (Figure 4C). Among these, the most influential variable was the first principal component of image features, followed by key inflammatory markers such as the C-reactive protein level and white blood cell count. Additional contributions from total bilirubin and liver-related enzyme levels further highlighted the complementary role of biochemical indicators.

**Figure 4:**
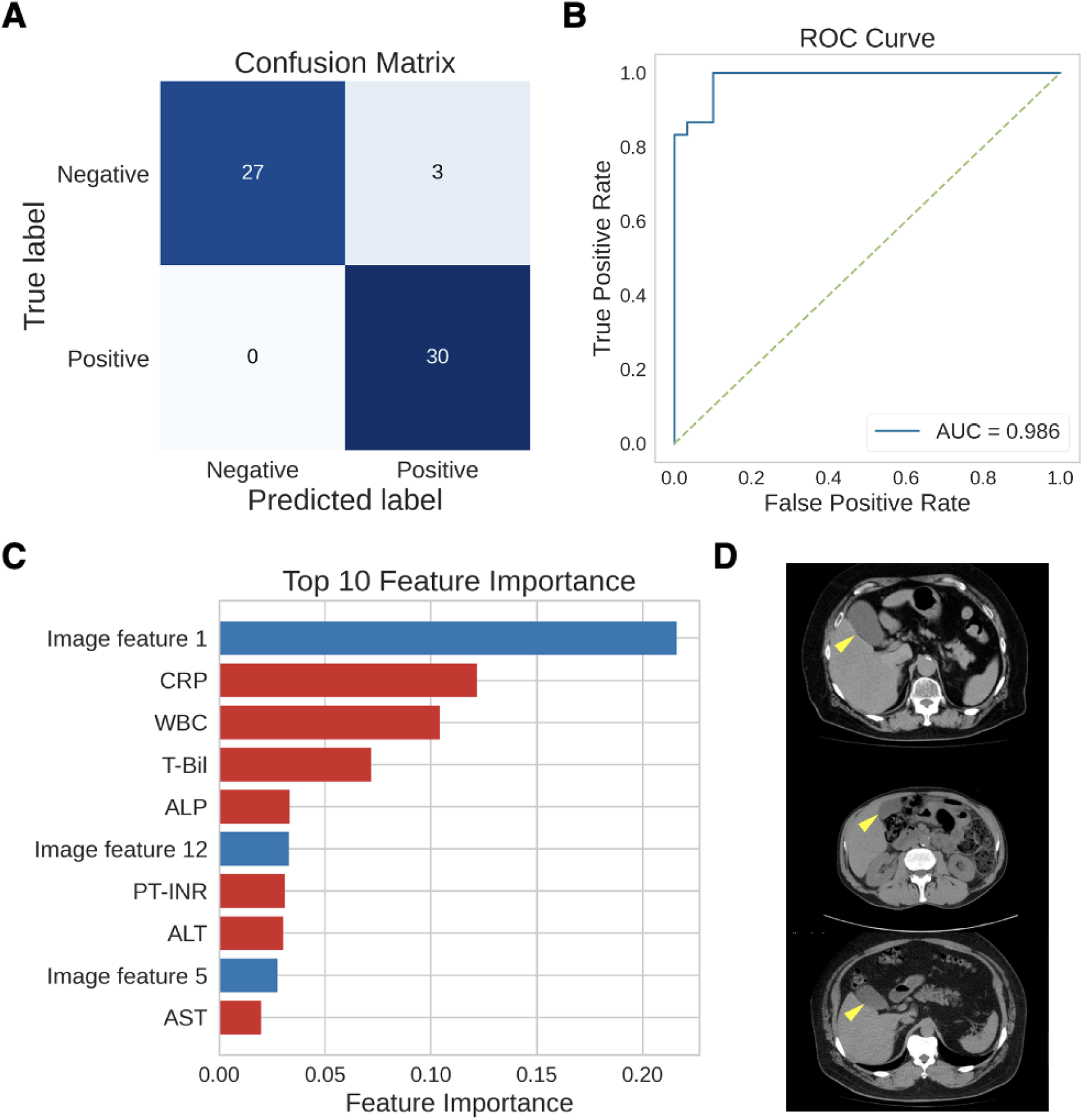
(A) Confusion matrix showing the true and predicted labels. (B) ROC curve evaluating the classification performance. The dashed line represents the performance of a random classifier. (C) Feature importance plot displaying the top 10 variables, including laboratory parameters and image-derived principal component analysis (PCA) features, contributing to the model’s predictions. (D) Three representative cases incorrectly classified as positive by the model. For each case, axial CT images are shown with the gallbladder indicated by yellow arrows.

## Discussion

In this study, we addressed the challenge of accurately diagnosing AC in emergency settings, where diagnostic decisions must balance accuracy with practical constraints such as invasiveness, cost, and time. These considerations highlight the clinical importance of diagnostic support based on non-contrast CT data. We developed three models: an Imaging Model, a Blood Model, and a Hybrid Model. The Hybrid Model achieved the best overall performance, underscoring the value of integrating imaging and clinical data.

The Imaging Model appeared to be influenced by morphologic features such as gallbladder distension, even without clear inflammatory findings. In contrast, the Blood Model was vulnerable in cases with attenuated inflammatory responses, such as chemotherapy-related cytopenia, potentially leading to false- negative predictions. Our results indicate complementary limitations of imaging and laboratory data, supporting the use of a hybrid approach to improve diagnostic robustness. Despite this advantage, the Hybrid Model still produced three false-positive cases. The upper and middle cases in Figure 4D were from patients with esophageal cancer prior to treatment initiation, during periods of poor oral intake. In fasting states, impaired gallbladder emptying can lead to physiologic distension, which may mimic imaging features of acute cholecystitis and contribute to false-positive classification (20,21). In contrast, the lower case in Figure 4D also involved a patient with esophageal cancer; however, oral intake was relatively preserved and CT was performed after a meal, a condition under which gallbladder distension is less likely. In this case, the false-positive classification was likely driven by discordant laboratory findings, including relatively elevated biliary enzyme levels (e.g., alkaline phosphatase and gamma-glutamyl transpeptidase; corresponding to the bottom case in Table 2) (22,23). Further refinement incorporating inflammation-specific imaging features and clinical context may improve specificity.

**Table 2.** Laboratory parameters in three misclassified cases.

| WBC | PLT | CRP | T-Bil | AST | ALT | γ GTP | ALP | Cre | PT-INR |
| --- | --- | --- | --- | --- | --- | --- | --- | --- | --- |
| [/μL] | [×10<sup>4</sup>/μL] | [mg/dL] | [mg/dL] | [U/L] | [U/L] | [U/L] | [U/L] | [mg/dL] |  |
| 8670 | 25 | 0.19 | 0.7 | 22 | 18 | 26 | 104 | 1.29 | 0.95 |
| 5260 | 19.4 | 0.34 | 1.1 | 19 | 11 | 26 | 58 | 0.95 | 0.95 |
| 7290 | 29.4 | 0.41 | 0.6 | 14 | 12 | 72 | 187 | 1.00 | 1.14 |

Data are presented as values for each case. WBC, white blood cell count; PLT, platelet count; CRP, C-reactive protein; T-Bil, total bilirubin; AST, aspartate aminotransferase; ALT, alanine aminotransferase; γGTP, gamma-glutamyl transferase; ALP, alkaline phosphatase; Cre, creatinine; PT-INR, prothrombin time–international normalized ratio

While point-of-care ultrasound remains widely used, its diagnostic performance is often influenced by operator expertise and availability, with a reported sensitivity and specificity of approximately 79.0% and 93.6%, respectively (15,24). To address these limitations, CT-based frameworks have been developed, with reported AUCs ranging from 0.7933 and to 0.8569 (10). In the present study, the Blood Model achieved comparable specificity (0.933) and precision (0.931) while improving recall (0.900 vs. 0.7867), supporting its potential clinical utility. This improved performance may be attributed, at least in part, to the use of HU values decomposed into RGB channels as model input. Notably, the Hybrid Model achieved a recall of 1.00, indicating no missed cases in this cohort, which may be advantageous in clinical settings where minimizing false negatives is critical. By reducing reliance on operator-dependent modalities, this approach may improve consistency in clinical decision-making and support triage, particularly in emergency settings where initial assessments are performed by non-specialists.

This study was subject to several limitations. First, the retrospective design may have introduced selection and information biases (25). Second, the heterogeneity in CT acquisition parameters and hardware across different centers may have influenced model generalizability, necessitating further external validation (26,27). Finally, the reference standard, derived from the Tokyo Guidelines 2018 criteria and composite clinical data, carries an inherent risk of label uncertainty. Future research should focus on prospective validation and the implementation of domain adaptation techniques to account for inter-institutional variability. Furthermore, ensuring transparency in artificial intelligence model development is paramount; subsequent reporting should adhere to the TRIPOD+AI (28), CLAIM 2024 (29), and STARD-AI (30) guidelines to ensure that these tools are both reproducible and ethically sound.

In conclusion, a deep learning-based hybrid model using non-contrast CT images showed potential as a diagnostic support tool for acute cholecystitis. In particular, it may serve as an objective and clinically implementable adjunct in emergency settings where junior residents and non-gastrointestinal specialists are involved in the initial assessment.

## Data Availability

The datasets generated and/or analyzed during the present study are available from the corresponding author upon reasonable request. All scripts used in this study are available on GitHub (https://github.com/RyosukeNomural/model_for_acute_cholecystitis).

## Declarations

## Ethical approval and consent to participate

This study was conducted in accordance with the Declaration of Helsinki. Given the retrospective nature of the study and the use of anonymized data, the requirement for informed consent was waived by the institutional ethics committee.

## Consent for publication

Not applicable.

## Availability of data and materials

The datasets used and/or analyzed during the current study are available from the corresponding author upon reasonable request.

## Competing interests

The authors declare that they have no competing interests.

## Funding

This study was supported by the 2024 “Clinical Research Grant for Young Physicians Engaged in Abdominal Emergency Care” from the Japanese Association for Abdominal Emergency Medicine and JH grant (2026-A-01). The funding body had no role in the study design; data collection, analysis, or interpretation; manuscript preparation; or the decision to submit for publication.

## Authors’ contributions

TH conceived and designed the study and drafted the manuscript. RT, Noriki Mitsui, YH, Ryoma Yokoi, Masashi Kuno, HH, MF, YS, IY, and RA contributed to data acquisition. RN, SAS, NS, KK, MO, HB, and Riu Yamashita contributed to data analysis, interpretation, and model development. Riu Yamashita critically revised the manuscript for important intellectual content. Nobuhisa Matsuhashi supervised the study and finalized the manuscript. All authors read and approved the final manuscript.

## Acknowledgments

We would like to thank Editage (www.editage.jp) for the English language editing. Computational analyses were performed on the KASHIWARP server at the National Cancer Center.

## Notes

### Competing Interest Statement

The authors have declared no competing interest.

### Author Declarations

The study protocol was approved by the Institutional Review Board of Gifu University Hospital (Approval No. 2024-007).

