## Supplemental_Files for "Development of a Deep Learning Model Integrating CT Images and Blood Data for the Diagnosis of Acute Cholecystitis"

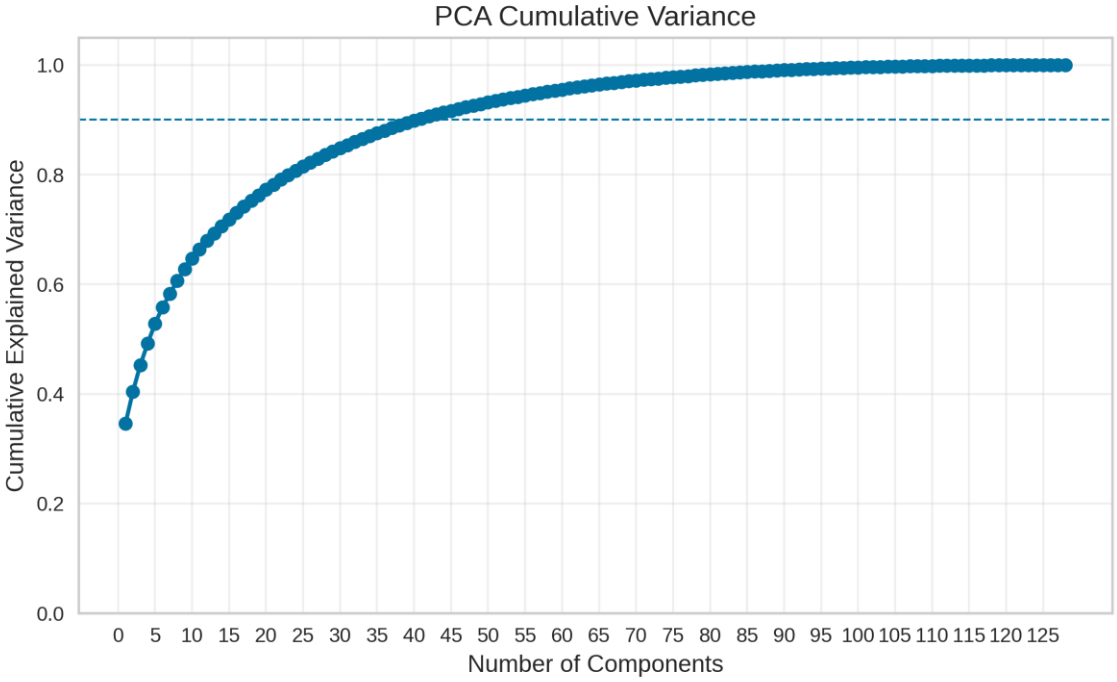


**Figure S1. Cumulative explained variance of principal component analysis (PCA) for image-derived features.**

The cumulative explained variance increased with the number of principal components, and 41 components were required to retain 90% of the variance. Accordingly, the first 41 principal components were used as image features in the hybrid model.

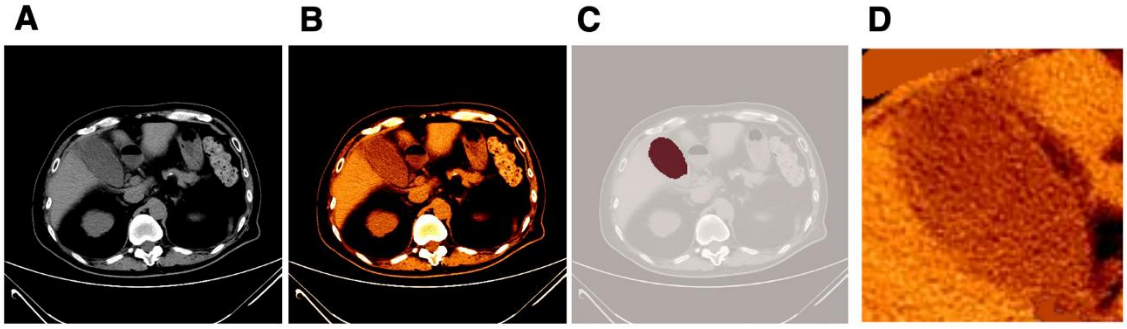


**Figure S2. Preprocessing pipeline for computed tomography (CT) image preparation.**

(A) Original CT image converted from DICOM to grayscale format. (B) Corresponding image converted to an RGB (three-channel) representation. (C) Automated gallbladder segmentation using TotalSegmentator, with the segmented region overlaid on the CT image.

(D) Cropped image centered on the segmented region with an additional margin, followed by correction of white saturation (blowout) artifacts.

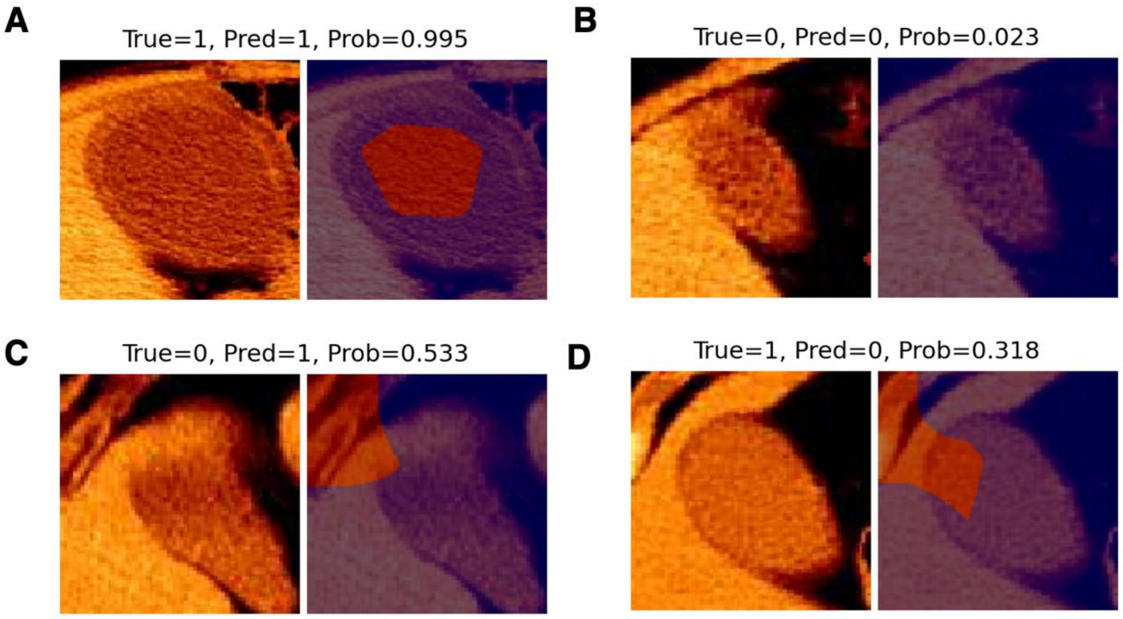


**Figure S3. Representative Gradient-weighted Class Activation Mapping (Grad-CAM) visualizations of the Imaging Model.**

(A–D) Representative cases showing input images (left) and corresponding Grad-CAM attention maps (right). Grad-CAM highlights regions contributing to the model's decision. Each panel includes the ground-truth label (True), model prediction (Pred), and predicted probability (Prob). (A) True-positive case. (B) True-negative case. (C) False-positive case. (D) False-negative case.

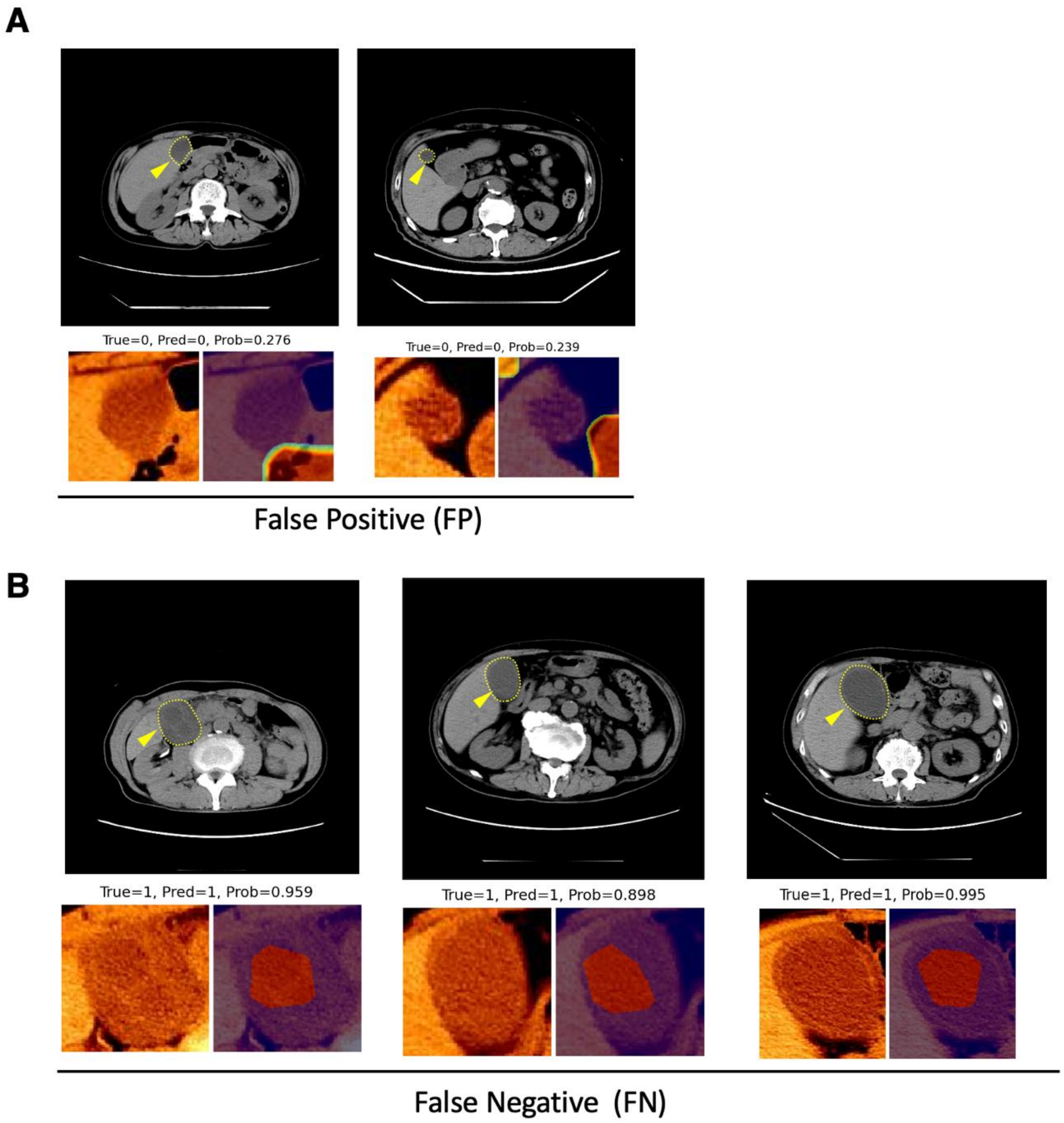


**Figure S4. Representative cases illustrating discordance between blood-based predictions and image-based features.**

(A) False positive cases from the Blood Model (True = 0, Pred = 1). Although classified as positive based on blood data, the corresponding computed tomography (CT) images and Gradient-weighted Class Activation Mapping (Grad-CAM) maps from the Imaging Model suggest limited or nonspecific imaging findings.

(B) False-negative cases from the Blood Model (True = 1, Pred = 0). Despite misclassification by the Blood Model, the Imaging Model highlights regions corresponding to the gallbladder, indicating imaging features consistent with disease.

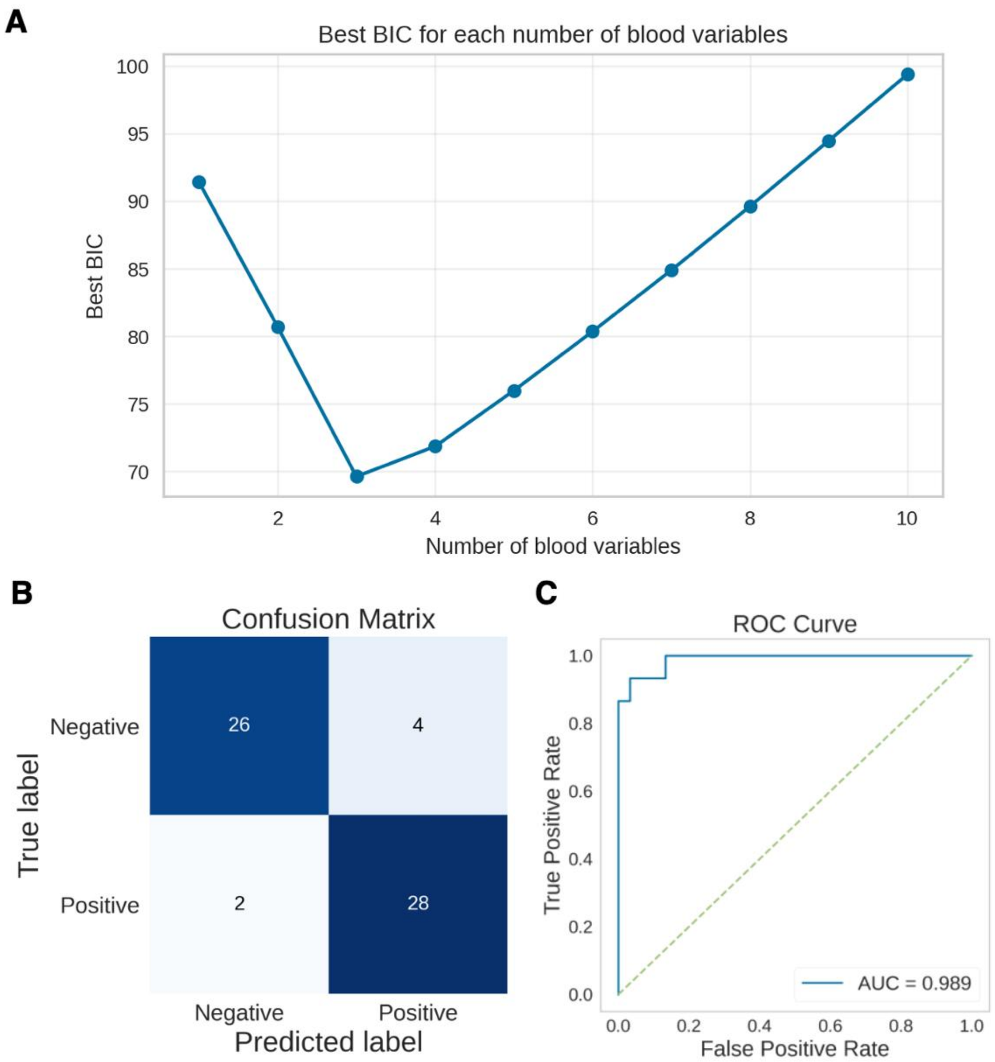


**Figure S5. Model optimization and performance of the Hybrid Model**

(A) Model selection for blood variables in the Hybrid Model based on the Bayesian Information Criterion (BIC). All possible combinations of blood variables (from 1 to 10 variables) were evaluated while keeping the image-derived features fixed. Variables were sequentially selected in the following order based on BIC minimization: CRP, ALP, WBC, ALT, γGTP, Cre, PLT, PT-INR, AST, and T-Bil. For each number of blood variables, the model with the lowest BIC was selected. The optimal model was achieved with three blood variables (CRP, ALP, WBC), which yielded the minimum BIC, indicating the best trade-off between model fit and complexity. (B) Confusion matrix of the hybrid model using the optimal combination of three blood variables. (C) Receiver operating characteristic (ROC) curve of the hybrid model with the optimal three blood variables.

Although the three-variable model achieved the lowest BIC, the model incorporating all blood variables demonstrated superior predictive performance and was therefore selected as the final model for the main analysis. CRP, C-reactive protein; ALP, alkaline phosphatase; WBC, white blood cell count; ALT, alanine aminotransferase; γGTP, gamma-glutamyl transpeptidase; Cre, creatinine; PLT, platelet count; PT-INR, prothrombin time–international normalized ratio; AST, aspartate aminotransferase; T-Bil, total bilirubin

| Label | WBC [ /µL] | PLT [$\times 10^4$/µL] | CRP [mg/dL] | T-Bil [mg/dL] | AST [U/L] | ALT [U/L] | γGTP [U/L] | ALP [U/L] | Cre [mg/dL] | PT-INR |
|---|---|---|---|---|---|---|---|---|---|---|
| FP | 6510 | 23.0 | 9.16 | 0.7 | 8 | 8 | 61 | 134 | 0.92 | 0.94 |
| FP | 8170 | 31.3 | 9.96 | 0.6 | 1 | 19 | 190 | 240 | 0.93 | 1.58 |
| FN | 5080 | 21.7 | 0.41 | 1.7 | 21 | 20 | 30 | 251 | 0.91 | 0.96 |
| FN | 2500 | 3.0 | 0.72 | 1.2 | 22 | 10 | 23 | 104 | 0.73 | 0.89 |
| FN | 9650 | 15.4 | 0.18 | 0.9 | 17 | 14 | 16 | 227 | 0.79 | 0.99 |

**Table S1. Laboratory parameters in five misclassified cases of the Blood Model.**

Blood test results for cases misclassified by the Blood Model, including two false-positive (FP) and three false-negative (FN) predictions. WBC, white blood cell count; PLT, platelet count; CRP, C-reactive protein; T-Bil, total bilirubin; AST, aspartate aminotransferase; ALT, alanine aminotransferase; γGTP, gamma-glutamyl transferase; ALP, alkaline phosphatase; Cre, creatinine; PT-INR, prothrombin time–international normalized ratio.
